# Immune protection against SARS-CoV-2 re-reinfection and immune imprinting

**DOI:** 10.1101/2022.08.23.22279026

**Authors:** Hiam Chemaitelly, Houssein H. Ayoub, Patrick Tang, Mohammad R. Hasan, Peter Coyle, Hadi M. Yassine, Hebah A. Al-Khatib, Maria K Smatti, Zaina Al-Kanaani, Einas Al-Kuwari, Andrew Jeremijenko, Anvar Hassan Kaleeckal, Ali Nizar Latif, Riyazuddin Mohammad Shaik, Hanan F. Abdul-Rahim, Gheyath K. Nasrallah, Mohamed Ghaith Al-Kuwari, Adeel A. Butt, Hamad Eid Al-Romaihi, Mohamed H. Al-Thani, Abdullatif Al-Khal, Roberto Bertollini, Laith J. Abu-Raddad

## Abstract

We investigated epidemiological evidence for immune imprinting by comparing incidence of re-reinfection in the national cohort of individuals with a documented Omicron (BA.1/BA.2) reinfection after a pre-Omicron primary infection (designated as the reinfection cohort), to incidence of reinfection in the national cohort of individuals with a documented Omicron (BA.1/BA.2) primary infection (designated as the primary-infection cohort). This was done using a matched, retrospective cohort study that emulated a randomized “target trial”. Vaccinated individuals were excluded. Associations were estimated using Cox proportional-hazard regression models. Cumulative incidence of infection was 1.1% (95% CI: 0.8-1.4%) for the reinfection cohort and 2.1% (95% CI: 1.8-2.3%) for the primary-infection cohort, 135 days after the start of follow-up. The adjusted hazard ratio (aHR) for infection was 0.52 (95% CI: 0.40-0.68), comparing incidence in the reinfection cohort to that in the primary-infection cohort. The aHR was 0.59 (95% CI: 0.40-0.85) in a subgroup analysis in which primary infection in the reinfection cohort was restricted to only the index virus or Alpha variant. In the first 70 days of follow-up, when incidence was dominated by BA.2, the aHR was 0.92 (95% CI: 0.51-1.65). However, cumulative incidence curves diverged when BA.4/BA.5 subvariants dominated incidence (aHR, 0.46 (95% CI: 0.34-0.62)). There was no evidence that immune imprinting compromises protection against Omicron subvariants. However, there was evidence that having two infections, one with a pre-Omicron variant followed by one with an Omicron subvariant, elicits stronger protection against future Omicron-subvariant reinfection than having had only one infection with an Omicron subvariant.

## Introduction

More than two years into the Coronavirus Disease 2019 (COVID-19) pandemic, the global population carries heterogenous immune histories derived from various exposures to infection, viral variants, and vaccination.^1^ Evidence at the level of binding and neutralizing antibodies, B cell, and T cell immunity suggests that previous severe acute respiratory syndrome coronavirus 2 (SARS-CoV-2) infection history can imprint a negative impact on subsequent protective immunity.^1^ In particular, immune response against Omicron (B.1.1.529) subvariants could be compromised by differential imprinting in those who had a prior infection with the index virus or Alpha (B.1.1.7) variant.^1^

We investigated epidemiological evidence for immune imprinting by comparing incidence of re-reinfection in the national cohort of individuals with a documented Omicron (B.1.1.529) (BA.1/BA.2^2^) reinfection after a pre-Omicron primary infection (designated as the reinfection cohort), to incidence of reinfection in the national cohort of individuals with a documented Omicron (BA.1/BA.2^2^) primary infection (designated as the primary-infection cohort). This was done using a matched, retrospective cohort study that emulated a randomized “target trial”.^3,4^

## Methods

### Study population and data sources

The study was conducted in the population of Qatar and analyzed COVID-19 data for laboratory testing, vaccination, hospitalization, and death, retrieved from the national digital-health information platform. Databases include all SARS-CoV-2-related data, with no missing information since pandemic onset, such as all polymerase chain reaction (PCR) tests, and starting from January 5, 2022, rapid antigen tests conducted at healthcare facilities.

Every PCR test (but not every rapid antigen test) conducted in Qatar is classified on the basis of symptoms and the reason for testing (clinical symptoms, contact tracing, surveys or random testing campaigns, individual requests, routine healthcare testing, pre-travel, at port of entry, or other). PCR and rapid antigen testing in Qatar is done at a mass scale, where a significant proportion of the population is tested every week.^5^ Most infections are diagnosed not because of appearance of symptoms, but because of routine testing.^5^ Qatar has unusually young, diverse demographics, in that only 9% of its residents are ≥50 years of age, and 89% are expatriates from over 150 countries.^6,7^ Qatar launched its COVID-19 vaccination program in December of 2020 using BNT162b2 and mRNA-1273 vaccines.^8^

Laboratory methods are in Supplementary Section S1. Classification of COVID-19 case severity (acute-care hospitalizations),^9^ criticality (intensive-care-unit hospitalizations),^9^ and fatality^10^ followed World Health Organization guidelines (Section S2). Further descriptions of the study population and these national databases were reported previously.^4,5,7,11,12^

### Study design

This national, matched, retrospective cohort study compared incidence of re-reinfection, irrespective of symptoms, in the national cohort of individuals with a documented Omicron-subvariant (BA.1/BA.2^2^) SARS-CoV-2 reinfection after an earlier pre-Omicron primary infection (the reinfection cohort), to incidence of reinfection in the national cohort of documented primary Omicron-subvariant (BA.1/BA.2^2^) infection (the primary-infection cohort).

Previous infections were classified as pre-Omicron versus Omicron previous infections, based on whether they occurred before or after the Omicron wave that started in Qatar on December 19, 2021.^12^ Incidence of non-Omicron variants has been limited since onset of the Omicron wave in Qatar.^4,12-14^

Documentation of infection in all cohorts was based on positive PCR or rapid antigen tests.

### Cohort eligibility, matching, and follow-up

Any individual with a documented reinfection between December 19, 2021 (onset of the Omicron wave in Qatar^2,4,12,13^) and August 15, 2022 was eligible for inclusion in the reinfection cohort, provided that the individual received no vaccination before the start of follow-up, set at 90 days after reinfection. Any individual with a documented primary infection between December 19, 2021 and August 15, 2022 was eligible for inclusion in the primary-infection cohort, provided that the individual received no vaccination before the start of follow-up. The primary study outcome was incidence of infection. The secondary outcome was incidence of severe, critical, or fatal COVID-19.

Individuals in the reinfection cohort were exact-matched in a one-to-three ratio by sex, 10-year age group, nationality, comorbidity count (none, 1 comorbidity, 2 comorbidities, 3 or more comorbidities), and calendar week of reinfection (Reinfection cohort)/calendar week of primary infection (Primary-infection cohort) to individuals in the primary-infection cohort, to control for differences in risk of SARS-CoV-2 infection in Qatar.^7,15-18^ Matching by these factors was shown previously to provide adequate control of differences in risk of infection.^5,8,19-21^ Matching was performed using an iterative process so that each individual in both cohorts was alive and unvaccinated at the start of follow-up.

SARS-CoV-2 reinfection is conventionally defined as a documented infection ≥90 days after an earlier infection, to avoid misclassification of prolonged PCR positivity as reinfection.^13,22,23^ Therefore, matched pairs were followed from the calendar day the individual in the reinfection cohort completed 90 days after the documented Omicron-subvariant reinfection.

For exchangeability,^4,24^ all members of matched pairs were censored on the earliest occurrence of first-dose vaccination of an individual in either cohorts. Individuals were followed up until the first of any of the following events: a documented SARS-CoV-2 infection, i.e., the first PCR-positive or rapid-antigen-positive test after the start of follow-up, regardless of symptoms, or first-dose vaccination (with matched pair censoring), or death, or end of study censoring.

### Statistical analysis

Eligible and matched cohorts were described using frequency distributions and measures of central tendency, and were compared using standardized mean differences (SMDs). An SMD ≤0.1 indicated adequate matching.^25^ Cumulative incidence of infection (defined as the proportion of individuals at risk, whose primary endpoint during follow-up was a re-reinfection for the reinfection cohort, or a reinfection for the primary-infection cohort) was estimated using the Kaplan–Meier estimator method.^26^ Incidence rate of infection in each cohort, defined as the number of identified infections divided by the number of person-weeks contributed by all individuals in the cohort, was estimated with its 95% confidence interval (CI) using a Poisson log-likelihood regression model with the Stata 17.0 *stptime* command.

The hazard ratio, comparing incidence of infection in both cohorts, and the corresponding 95% CI, were calculated using Cox regression adjusted for matching factors with the Stata 17.0 *stcox* command. Schoenfeld residuals and log-log plots for survival curves were used to test the proportional-hazards assumption and to investigate its adequacy. 95% CIs were not adjusted for multiplicity; thus, they should not be used to infer definitive differences between cohorts. Interactions were not considered.

A subgroup analysis was conducted to estimate adjusted hazard ratios by time since reinfection. This was done using separate Cox regressions with “failures” restricted to specific time intervals. Another subgroup analysis was conducted to investigate immune protection in the situation that the primary infection in the reinfection cohort was restricted to only the index virus or the Alpha variant, out of specific relevance to effect of immune imprinting.^1^ Sensitivity analysis adjusting the hazard ratio by the ratio of testing frequencies between cohorts was also performed. Statistical analyses were conducted using Stata/SE version 17.0 (Stata Corporation, College Station, TX, USA).

### Oversight

Hamad Medical Corporation and Weill Cornell Medicine-Qatar Institutional Review Boards approved this retrospective study with a waiver of informed consent. The study was reported following the Strengthening the Reporting of Observational Studies in Epidemiology (STROBE) guidelines. The STROBE checklist is found in Table S1.

## Results

Figure 1 shows the population selection process and Table 2 describes baseline characteristics of the full and matched cohorts. The matched cohorts included 7,873 individuals in the reinfection cohort and 22,349 individuals in the primary-infection cohort. The study is representative of the unvaccinated population of Qatar.

**Table 2.**
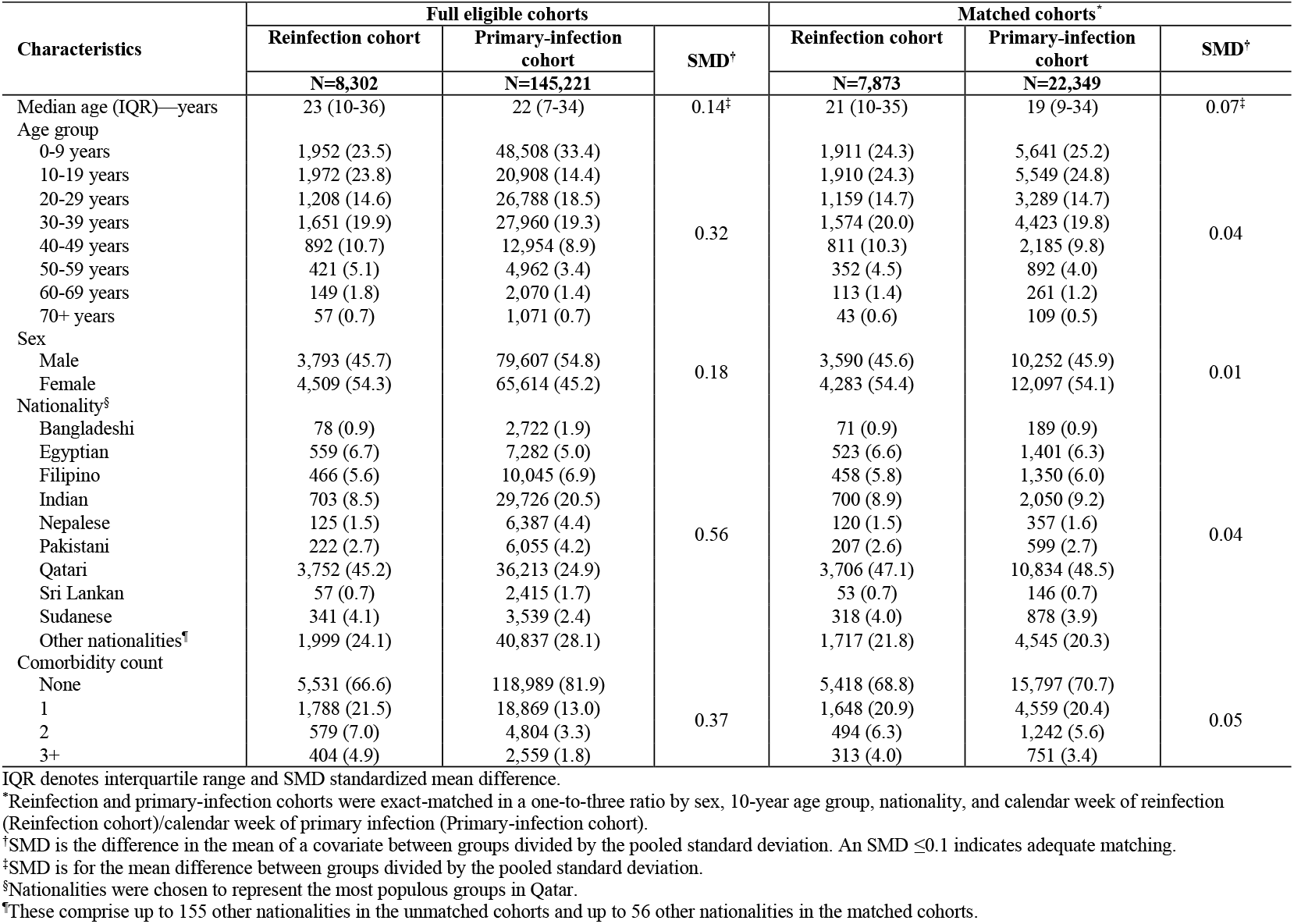
Baseline characteristics of the eligible and matched cohorts.

**Figure 1.**
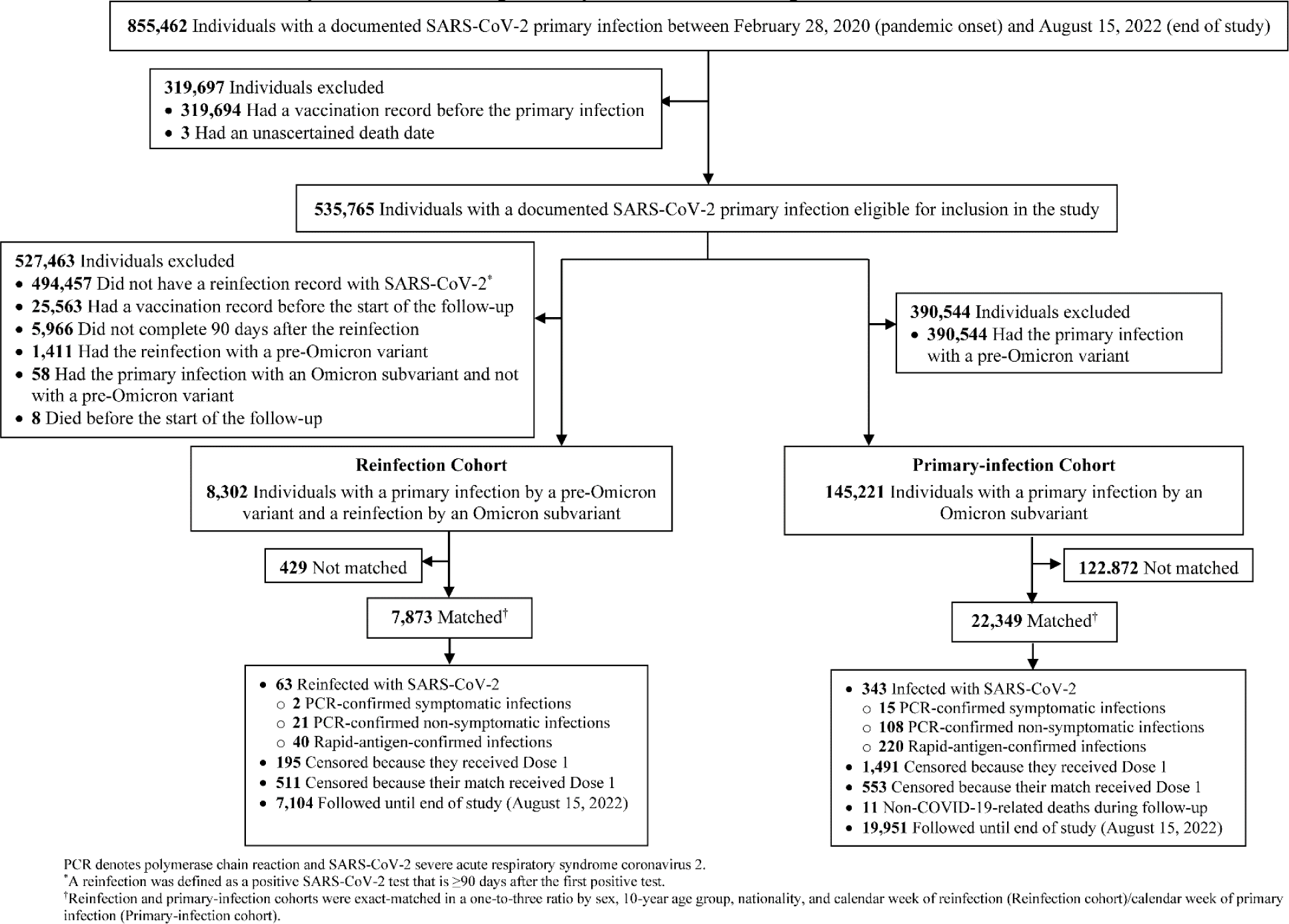
Flowchart describing the population selection process for investigating immune protection against reinfection among those who were infected by an Omicron subvariant compared to protection among those who were infected by an Omicron subvariant, but additionally had an earlier primary infection with a pre-Omicron variant.

There were 63 re-reinfections in the reinfection cohort and 343 reinfections in the primary-infection cohort during follow-up, none of which progressed to severe, critical, or fatal COVID-19 (Figure 1). Cumulative incidence of infection was 1.1% (95% CI: 0.8-1.4%) for the reinfection cohort and 2.1% (95% CI: 1.8-2.3%) for the primary-infection cohort, 135 days after the start of follow-up (Figure 2A).

**Figure 2.**
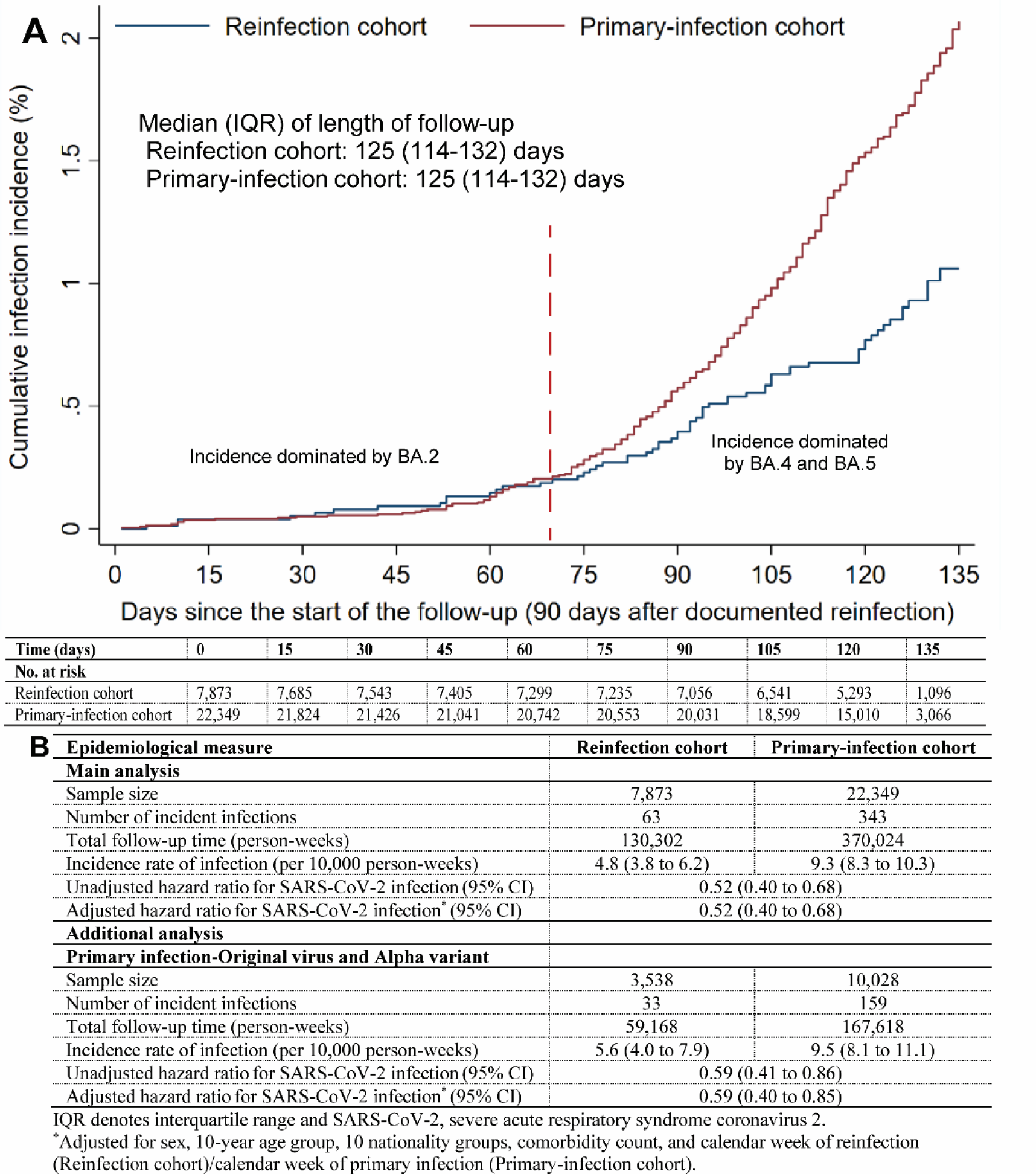
A) Cumulative incidence of reinfection and B) hazard ratios for the incidence of SARS-CoV-2 reinfection in the Reinfection and Primary-infection cohorts. This cohort study was conducted in the population of Qatar between December 19, 2021 and August 15, 2022.

The adjusted hazard ratio (aHR) for infection was 0.52 (95% CI: 0.40-0.68), comparing incidence in the reinfection cohort to that in the primary-infection cohort. The aHR was 0.59 (95% CI: 0.40-0.85) in the analysis in which primary infection in the reinfection cohort was restricted to only the index virus or Alpha variant (Figure 2B).

In the first 70 days of follow-up, when incidence was dominated by the BA.2 subvariant,^2,12^ the aHR was 0.92 (95% CI: 0.51-1.65). However, cumulative incidence curves diverged when BA.4/BA.5 subvariants were introduced and dominated incidence^14^ (aHR, 0.46 (95% CI: 0.34-0.62); Figure 2A).

Differences in testing frequency existed between the followed cohorts, but these were small. The proportion of individuals who had a SARS-CoV-2 test during follow-up was 45.3% for the reinfection cohort and 38.4% for the primary-infection cohort. The testing frequency was 0.79 and 0.64 tests per person, respectively. Adjusting the hazard ratio estimate in a sensitivity analysis by the ratio of testing frequencies between cohorts yielded an adjusted hazard ratio of 0.42 (95% CI: 0.32-0.55), confirming study results.

## Discussion

There was no evidence that immune imprinting compromises protection against Omicron subvariants. However, there was evidence that having two infections, one with a pre-Omicron variant followed by one with an Omicron subvariant, elicits stronger protection against future Omicron-subvariant reinfection than having had only one infection with an Omicron subvariant. The differences in immune protection emerged when BA.4/BA.5 dominated incidence, perhaps suggesting that the earlier pre-Omicron infection may have contributed to broadening the immune response against a future reinfection challenge.

## Limitations

We investigated incidence of documented infections, but other infections may have occurred and gone undocumented. Undocumented infections confer immunity or boost existing immunity, thereby perhaps affecting the estimates. With Qatar’s young population and the young age of those that remained unvaccinated in our population, our findings may not be generalizable to older individuals or to other countries where elderly citizens constitute a larger proportion of the total population.

Depletion of the reinfection cohort by COVID-19 mortality at time of the primary infection may have biased this cohort towards healthier individuals with stronger immune responses. However, COVID-19 mortality has been low in Qatar’s predominantly young population,^7,27^ totaling 681 COVID-19 deaths (<0.1% of primary infections) up to August 15, 2022. A survival effect seems unlikely to explain or appreciably affect study findings.

As an observational study, investigated cohorts were neither blinded nor randomized, so unmeasured or uncontrolled confounding cannot be excluded. While matching was done for sex, age, nationality, comorbidity count, and calendar week of Omicron-subvariant infection, this was not possible for other factors such as geography or occupation, as such data were unavailable. However, Qatar is essentially a city state and infection incidence was broadly distributed across neighborhoods. Nearly 90% of Qatar’s population are expatriates from over 150 countries, coming here because of employment.^7^ Most are craft and manual workers working in development projects.^7^ Nationality, age, and sex provide a powerful proxy for socio-economic status in this country.^7,15-18^ Nationality alone is strongly associated with occupation.^7,16-18^

Matching was done to control for factors that affect infection exposure in Qatar.^7,15-18^ The matching prescription had already been investigated in previous studies of different epidemiologic designs, and using control groups to test for null effects.^5,8,19-21^ These control groups included unvaccinated cohorts versus vaccinated cohorts within two weeks of the first dose,^5,19-21^ when vaccine protection is negligible,^28^ and mRNA-1273-versus BNT162b2-vaccinated cohorts, also in the first two weeks after the first dose.^8^ These studies have shown that this prescription provides adequate control of the differences in infection exposure.^5,8,19-21^ The present study analyses were implemented on Qatar’s total population, thus perhaps minimizing the likelihood of bias.

## Data Availability

The dataset of this study is a property of the Qatar Ministry of Public Health that was provided to the researchers through a restricted-access agreement that prevents sharing the dataset with a third party or publicly. Future access to this dataset can be considered through a direct application for data access to Her Excellency the Minister of Public Health (https://www.moph.gov.qa/english/Pages/default.aspx). Aggregate data are available within the manuscript.

## Acknowledgements

We acknowledge the many dedicated individuals at Hamad Medical Corporation, the Ministry of Public Health, the Primary Health Care Corporation, the Qatar Biobank, Sidra Medicine, and Weill Cornell Medicine – Qatar for their diligent efforts and contributions to make this study possible.

The authors are grateful for support from the Biomedical Research Program and the Biostatistics, Epidemiology, and Biomathematics Research Core, both at Weill Cornell Medicine-Qatar, as well as for support provided by the Ministry of Public Health, Hamad Medical Corporation, and Sidra Medicine. The authors are also grateful for the Qatar Genome Programme and Qatar University Biomedical Research Center for institutional support for the reagents needed for the viral genome sequencing. Statements made herein are solely the responsibility of the authors.

The funders of the study had no role in study design, data collection, data analysis, data interpretation, or writing of the article.

## Author contributions

HC co-designed the study, performed the statistical analyses, and co-wrote the first draft of the article. LJA conceived and co-designed the study, led the statistical analyses, and co-wrote the first draft of the article. PVC designed mass PCR testing to allow routine capture of SGTF variants. PT and MRH conducted the multiplex, RT-qPCR variant screening and viral genome sequencing. HY, HAK, and MKS conducted viral genome sequencing. All authors contributed to data collection and acquisition, database development, discussion and interpretation of the results, and to the writing of the manuscript. All authors have read and approved the final manuscript.

## Supplementary Appendix

### Section S1. Laboratory methods

#### Real-time reverse-transcription polymerase chain reaction testing

Nasopharyngeal and/or oropharyngeal swabs were collected for polymerase chain reaction (PCR) testing and placed in Universal Transport Medium (UTM). Aliquots of UTM were: 1) extracted on KingFisher Flex (Thermo Fisher Scientific, USA), MGISP-960 (MGI, China), or ExiPrep 96 Lite (Bioneer, South Korea) followed by testing with real-time reverse-transcription PCR (RT-qPCR) using TaqPath COVID-19 Combo Kits (Thermo Fisher Scientific, USA) on an ABI 7500 FAST (Thermo Fisher Scientific, USA); 2) tested directly on the Cepheid GeneXpert system using the Xpert Xpress SARS-CoV-2 (Cepheid, USA); or 3) loaded directly into a Roche cobas 6800 system and assayed with the cobas SARS-CoV-2 Test (Roche, Switzerland). The first assay targets the viral S, N, and ORF1ab gene regions. The second targets the viral N and E-gene regions, and the third targets the ORF1ab and E-gene regions.

All PCR testing was conducted at the Hamad Medical Corporation Central Laboratory or Sidra Medicine Laboratory, following standardized protocols.

#### Rapid antigen testing

Severe acute respiratory syndrome coronavirus 2 (SARS-CoV-2) antigen tests were performed on nasopharyngeal swabs using one of the following lateral flow antigen tests: Panbio COVID-19 Ag Rapid Test Device (Abbott, USA); SARS-CoV-2 Rapid Antigen Test (Roche, Switzerland); Standard Q COVID-19 Antigen Test (SD Biosensor, Korea); or CareStart COVID-19 Antigen Test (Access Bio, USA). All antigen tests were performed point-of-care according to each manufacturer’s instructions at public or private hospitals and clinics throughout Qatar with prior authorization and training by the Ministry of Public Health (MOPH). Antigen test results were electronically reported to the MOPH in real time using the Antigen Test Management System which is integrated with the national Coronavirus Disease 2019 (COVID-19) database.

#### Viral genome sequencing and classification of infections by variant type

Surveillance for SARS-CoV-2 variants in Qatar is based on viral genome sequencing and multiplex RT-qPCR variant screening^1^ of random positive clinical samples,^2-7^ complemented by deep sequencing of wastewater samples.^4,8,9^ Further details on the viral genome sequencing and multiplex RT-qPCR variant screening throughout the SARS-CoV-2 waves in Qatar can be found in previous publications.^2-7,10-16^

### Section S2. COVID-19 severity, criticality, and fatality classification

Classification of COVID-19 case severity (acute-care hospitalizations),^17^ criticality (intensive-care-unit hospitalizations),^17^ and fatality^18^ followed World Health Organization (WHO) guidelines. Assessments were made by trained medical personnel independent of study investigators and using individual chart reviews, as part of a national protocol applied to every hospitalized COVID-19 patient. Each hospitalized COVID-19 patient underwent an infection severity assessment every three days until discharge or death. We classified individuals who progressed to severe, critical, or fatal COVID-19 between the time of the documented infection and the end of the study based on their worst outcome, starting with death,^18^ followed by critical disease,^17^ and then severe disease.^17^

Severe COVID-19 disease was defined per WHO classification as a SARS-CoV-2 infected person with “oxygen saturation of <90% on room air, and/or respiratory rate of >30 breaths/minute in adults and children >5 years old (or ≥60 breaths/minute in children <2 months old or ≥50 breaths/minute in children 2-11 months old or ≥40 breaths/minute in children 1–5 years old), and/or signs of severe respiratory distress (accessory muscle use and inability to complete full sentences, and, in children, very severe chest wall indrawing, grunting, central cyanosis, or presence of any other general danger signs)”.^17^ Detailed WHO criteria for classifying SARS-CoV-2 infection severity can be found in the WHO technical report.^17^

Critical COVID-19 disease was defined per WHO classification as a SARS-CoV-2 infected person with “acute respiratory distress syndrome, sepsis, septic shock, or other conditions that would normally require the provision of life sustaining therapies such as mechanical ventilation (invasive or non-invasive) or vasopressor therapy”.^17^ Detailed WHO criteria for classifying SARS-CoV-2 infection criticality can be found in the WHO technical report.^17^

COVID-19 death was defined per WHO classification as “a death resulting from a clinically compatible illness, in a probable or confirmed COVID-19 case, unless there is a clear alternative cause of death that cannot be related to COVID-19 disease (e.g. trauma). There should be no period of complete recovery from COVID-19 between illness and death. A death due to COVID-19 may not be attributed to another disease (e.g. cancer) and should be counted independently of preexisting conditions that are suspected of triggering a severe course of COVID-19”. Detailed WHO criteria for classifying COVID-19 death can be found in the WHO technical report.^18^

**Table S1.** STROBE checklist for cohort studies.

|  | Item No | Recommendation | Main Text page |
| --- | --- | --- | --- |
| Title and abstract | 1 | (a) Indicate the study’s design with a commonly used term in the title or the abstract | Abstract |
|  |  | (b) Provide in the abstract an informative and balanced summary of what was done and what was found | Not applicable |
| Introduction |  |  |  |
| Background/rationale | 2 | Explain the scientific background and rationale for the investigation being reported | Introduction |
| Objectives | 3 | State specific objectives, including any prespecified hypotheses | Introduction |
| Methods |  |  |  |
| Study design | 4 | Present key elements of study design early in the paper | Methods (‘Study design’ & ‘Cohort eligibility, matching, and follow-up’) |
| Setting | 5 | Describe the setting, locations, and relevant dates, including periods of recruitment, exposure, follow-up, and data collection | Methods (‘Study population and data sources’, ‘Study design’ & ‘Cohort eligibility, matching, and follow-up’) |
| Participants | 6 | (a) Give the eligibility criteria, and the sources and methods of selection of participants. Describe methods of follow-up<br>(b) For matched studies, give matching criteria and number of exposed and unexposed | Methods (‘Study design’ & ‘Cohort eligibility, matching, and follow-up’) & Figure 1 |
| Variables | 7 | Clearly define all outcomes, exposures, predictors, potential confounders, and effect modifiers. Give diagnostic criteria, if applicable | Methods, Figure 1, & Table 2 |
| Data sources/ measurement | 8* | For each variable of interest, give sources of data and details of methods of assessment (measurement). Describe comparability of assessment methods if there is more than one group | Methods & Table 2 |
| Bias | 9 | Describe any efforts to address potential sources of bias | Methods (‘Cohort eligibility, matching, and follow-up’) |
| Study size | 10 | Explain how the study size was arrived at | Figure 1 |
| Quantitative variables | 11 | Explain how quantitative variables were handled in the analyses. If applicable, describe which groupings were chosen and why | Methods (‘Cohort eligibility, matching, and follow-up’) & Table 2 |
| Statistical methods | 12 | (a) Describe all statistical methods, including those used to control for confounding | Methods (‘Statistical analysis’) |
|  |  | (b) Describe any methods used to examine subgroups and interactions | Methods (‘Statistical analysis’) |
|  |  | (c) Explain how missing data were addressed | Not applicable, see Methods (‘Study population and data sources’) |
|  |  | (d) If applicable, explain how loss to follow-up was addressed | Not applicable, see Methods (‘Study population and data sources’) |
|  |  | (e) Describe any sensitivity analyses | Methods (‘Statistical analysis’) |
| Results |  |  |  |
| Participants | 13* | (a) Report numbers of individuals at each stage of study—eg numbers potentially eligible, examined for eligibility, confirmed eligible, included in the study, completing follow-up, and analysed<br>(b) Give reasons for non-participation at each stage<br>(c) Consider use of a flow diagram | Results, & Figure 1 |
| Descriptive data | 14 | (a) Give characteristics of study participants (eg demographic, clinical, social) and information on exposures and potential confounders | Table 2 |
|  |  | (b) Indicate number of participants with missing data for each variable of interest | Not applicable, see Methods ('Study population and data sources') |
|  |  | (c) Summarise follow-up time (eg, average and total amount) | Figure 2 |
| Outcome data | 15 | Report numbers of outcome events or summary measures over time | Figures 1 & 2 |
| Main results | 16 | (a) Give unadjusted estimates and, if applicable, confounder-adjusted estimates and their precision (eg, 95% confidence interval). Make clear which confounders were adjusted for and why they were included | Results & Figure 2 |
|  |  | (b) Report category boundaries when continuous variables were categorized | Table 2 |
|  |  | (c) If relevant, consider translating estimates of relative risk into absolute risk for a meaningful time period | Not applicable |
| Other analyses | 17 | Report other analyses done—eg analyses of subgroups and interactions, and sensitivity analyses | Results & Figure 2 |
| Discussion |  |  |  |
| Key results | 18 | Summarise key results with reference to study objectives | Discussion, paragraph 1 |
| Limitations | 19 | Discuss limitations of the study, taking into account sources of potential bias or imprecision. Discuss both direction and magnitude of any potential bias | Discussion ('Limitations') |
| Interpretation | 20 | Give a cautious overall interpretation of results considering objectives, limitations, multiplicity of analyses, results from similar studies, and other relevant evidence | Discussion, paragraph 1 |
| Generalisability | 21 | Discuss the generalisability (external validity) of the study results | Discussion ('Limitations') |
| Other information |  |  |  |
| Funding | 22 | Give the source of funding and the role of the funders for the present study and, if applicable, for the original study on which the present article is based | Acknowledgements |

